# Inhaled corticosteroid use in COVID-19

**DOI:** 10.1101/2020.09.03.20187278

**Authors:** Anders Husby, Anton Pottegård, Anders Hviid

## Abstract

**Background:** Recent evidence has established a beneficial effect of systemic corticosteroids for treatment of moderate-to-severe COVID-19. However, it is unknown if inhaled corticosteroid use is associated with reduced morbidity of the disease.

**Methods:** In a nationwide cohort of hospitalized SARS-CoV-2 test-positive individuals in Denmark, we estimated the 30-day hazard ratio of intensive care unit (ICU) admission or death among users of inhaled corticosteroids (ICS) compared with users of non-ICS inhalers (β_2_-agonist/muscarinic-antagonists), or non-users of ICS, with Cox regression adjusted for age, sex, and other confounders. We repeated these analyses among influenza test-positive patients during 2010–2018.

**Results:** Among 2,180 hospitalized SARS-CoV-2 patients, 282 were admitted to ICU and 421 died within 30 days. ICS use was associated with a hazard ratio of 1.25 (95% CI [CI], 0.60 to 2.61) for ICU admission and 0.84 (95% CI, 0.54 to 1.31) for death compared with non-ICS inhaler use. Compared with no ICS use, the hazard ratio of ICU admission or death was 1.22 (95% CI, 0.77 to 1.94) and 1.05 (95% CI, 0.75 to 1.47), respectively. Among 10,279 hospitalized influenza patients, the hazard ratios were 1.43 (95% CI, 0.89 to 2.30) and 1.11 (95% CI, 0.85 to 1.46) for ICU admission, and 0.80 (95% CI, 0.63 to 1.01) and 1.03 (95% CI, 0.87 to 1.22) for death compared with non-ICS inhaler use and no ICS use, respectively.

**Conclusions:** Our results do not support an effect of inhaled corticosteroid use on COVID-19 morbidity, however we can only rule out moderate-to-large reduced or increased risks.

## BACKGROUND

Infection with the novel coronavirus SARS-CoV-2, the causative agent of the COVID-19 pandemic, does not yet have any specific prophylaxis and only limited treatment options (1–3). Studies report that SARS-CoV-2 infection often leads to severe airway inflammation(4) and the RECOVERY-trial suggests a substantial beneficial effect of systemic treatment with the corticosteroid dexamethasone in hospitalized COVID-19 patients requiring nasal oxygen or mechanical ventilation(3). On the other hand, a tendency towards an adverse effect of per oral dexamethasone for patients not requiring oxygen was found in the trial. Nevertheless, the role of inhaled corticosteroids in morbidity of COVID-19 is unknown. Adding to the uncertainty, pre-clinical studies suggests inhaled corticosteroids downregulate the SARS-CoV-2 receptors ACE2/TMPRSS2(5) and inhibit SARS-CoV-2 replication(6), while there is evidence of more severe disease in COPD patients(7,8).

Using the unique Danish nationwide registers on prescription drug use, laboratory-confirmed infectious disease, and hospitalizations, we present a nationwide cohort study of inhaled corticosteroid use and COVID-19 outcomes. To aid in the interpretation of the effects of inhaled corticosteroid use on COVID-19 morbidity in a real-world setting, we conducted a comparison analysis of the effect of inhaled corticosteroid use on influenza morbidity during the 2010–2018 influenza seasons.

## METHODS

### Materials

The Danish Civil Registration System (CRS) allows individual-level linkage of information from national health registers, in addition to providing demographic information on the Danish population(9). Information on PCR tests for SARS-CoV-2 and influenza infection is available through MiBA, the Danish Microbiology Database, which includes all microbiological test results in Denmark, starting from 2010(10–12). The Danish National Patient Register covers information on hospital admission, admission to intensive care units (ICU), use of mechanical ventilation, and diagnostic codes to identify underlying comorbidities(13). The Danish National Prescription Registry, which contains individual-level information on all filled prescriptions in Denmark, provides information on pharmaceutical exposures of interest(14). The Cause of Death Register includes information on all registered deaths in Denmark(15).

### Study population

All hospitalized individuals aged 40 years or older in Denmark with a positive SARS-CoV-2 PCR test up to July 16, 2020, were included in our COVID-19 cohort from the date of testing or hospitalization, whichever came latest. The COVID-19 cohort was followed up for ICU admission, death, or loss to follow-up within 30 days from cohort entry. Individuals who tested PCR-positive for influenza during 2010–2018 were included in an equivalent influenza cohort from the date of testing or hospitalization, whichever came latest, and followed up for ICU admission, death, or loss to follow-up within 30 days from cohort entry. For sensitivity analyses, we also constructed a nationwide cohort of all individuals aged 40 years or older who tested positive for SARS-CoV-2 while out-of-hospital. This cohort was followed up for hospitalization, loss to follow-up, or death within 30 days from the test date. In addition, we constructed a cohort of SARS-CoV-2 test-positive ICU-patients who were followed up for death within 30 days from admission to ICU.

### Study variables

Exposure groups were categorized as 1) individuals with inhaled corticosteroid (ICS) use, defined as one or more filled prescriptions of inhaled corticosteroids within the last six months, with or without simultaneous filled prescriptions for other inhaled pharmaceuticals (i.e. β_2_-receptor agonist and/or muscarinic receptor antagonists), or use of combinatory inhalers (e.g. combined ICS and β_2_-receptor agonist inhaler), 2) individuals with β_2_-receptor agonist and/or muscarinic receptor antagonists use defined as one or more filled prescriptions within the last six months, but not ICS use, and 3) individuals without ICS use. Prescriptions two weeks prior to a positive test were omitted when allocating study participants to exposure groups. Information on other covariates of interest was defined by relevant pharmaceutical, demographic, and diagnostic codes (see Table S1 for detailed description).

### Outcomes

Information on date of admission to ICU and mechanical ventilation, respectively, was acquired from the Danish National Patient Register. Information on date of death was acquired from the Cause of Death Register.

### Statistical analysis

Our main analysis was conducted among hospitalized individuals who tested positive for SARS-CoV-2 (in 2020) and influenza (in 2010–2018), respectively. We followed participants for 30 days from the date of study entry until either ICU admission, death, or loss to follow-up from other causes. We used Cox proportional hazards regression to estimate the hazard ratios of death and ICU admission comparing exposure groups. We estimated 30-day cumulative hazards according to exposure status taking competing risks into account using the Nelson-Aalen estimator. In the Cox models, we took potential confounders into account through direct propensity score adjustment, in addition to age, sex, β2-receptor agonist use, and muscarinic receptor antagonist use. We considered the following covariates in the propensity score: atrial fibrillation, dementia, heart failure, hypertension, inflammatory bowel disease, malignancy, renal failure, Charlson Comorbidity index score, number of filled prescription within 90 days, and per oral corticosteroid use. Covariate status was ascertained 6 months before study entry (before exposure ascertainment). Propensity scores were estimated using logistic regression of probability of exposure on the above-mentioned covariates as main effects. We estimated separate propensity scores for each exposure group of interest (distribution of propensity scores by exposure group can be seen in Supplementary Figures S1-S4 for the COVID-19 cohort and Figures S5-S8 for the influenza cohort).

## RESULTS

Our COVID-19 cohort included 2,180 individuals who had been hospitalized with a positive SARS-CoV-2 test up to and including July 16, 2020, while our influenza cohort included 10,279 individuals hospitalized with a positive influenza test during 2010–2018. The cohort of COVID-19 patients had a noticeably lower frequency of comorbidities compared with the cohort of influenza patients, except a marginally higher prevalence of dementia in the COVID-19 cohort (Table 1). Furthermore, COVID-19 patients were slightly younger and had fewer filled prescriptions within the last 90 days compared with influenza patients. Among COVID-19 patients, 5.7% were registered with a hospital diagnosis for asthma and 9.6% with a chronic pulmonary disease (incl. COPD) diagnosis (Table 1). While 16.7% had filled a prescription for any inhaled pharmaceutical within the last six months (defined as usage), 10.4%, 8.7%, 8.5%, 1.1%, and 6.3% had used inhaled pharmaceutical containing corticosteroids (ICS), inhaled short-acting β_2_-agonist (SABA), inhaled long-acting β_2_-agonist (LABA), inhaled short-acting muscarinic antagonist (SAMA), or inhaled long-acting muscarinic antagonist (LAMA), respectively. Among influenza patients, 5.8% and 13.3% were registered with an asthma and a chronic pulmonary disease diagnosis, respectively. Any inhaled pharmaceutical use (26.9%) comprised ICS (18.7%), SABA (16.5%), LABA (13.3%), SAMA (2.3%) and LAMA (13.9%) use, respectively.

**Table 1.**
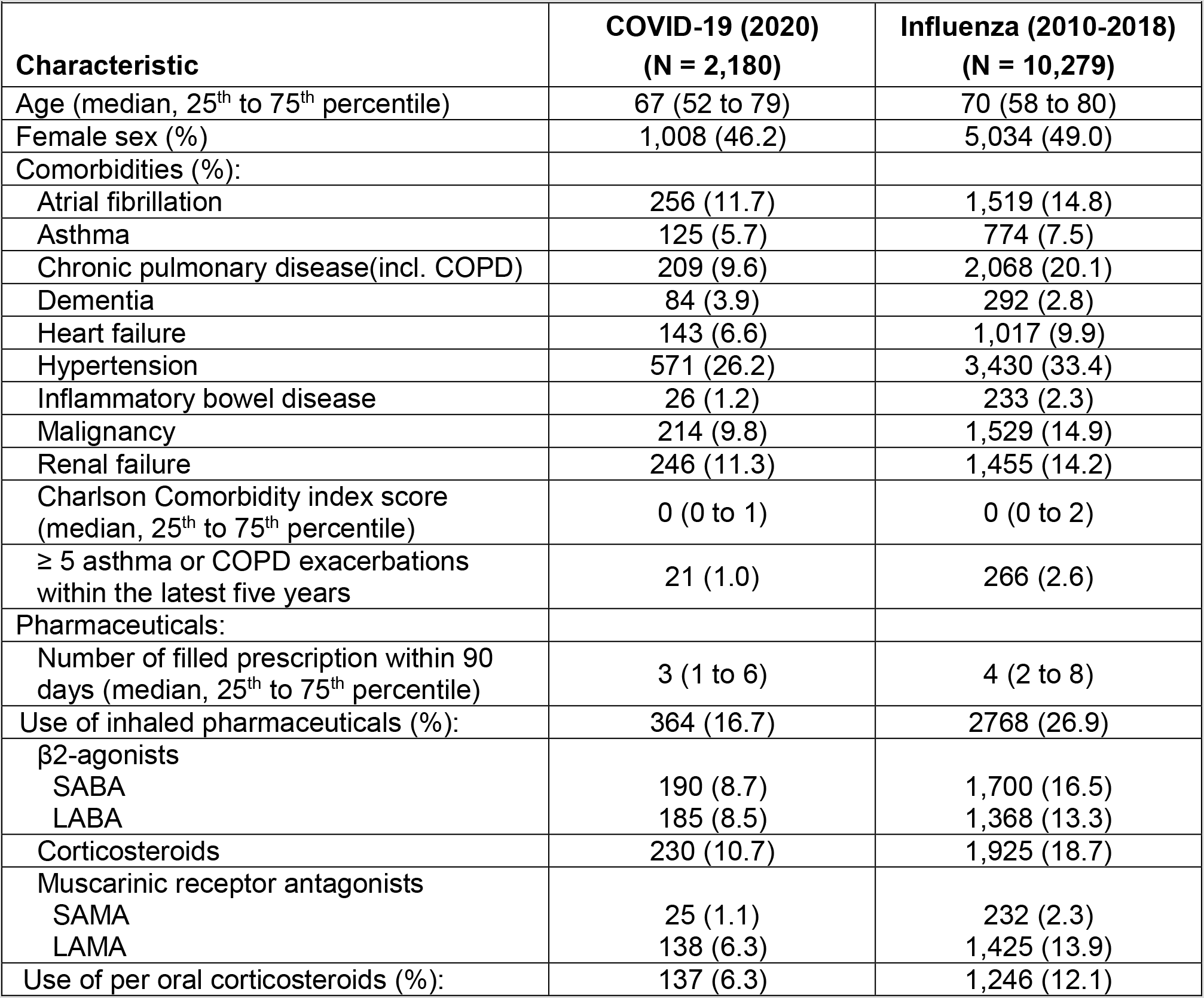
Baseline characteristics (age, sex, comorbidities, and prescription drug use) of the Danish cohorts of hospitalized individuals tested positive for SARS-CoV-2 during the COVID-19 epidemic, and individuals tested positive for influenza during 2010-2018, respectively.

In our main analysis of COVID-19 patients, the 30-day hazard of admission to intensive care unit (ICU) was similar among ICS users compared with users of other inhaled pharmaceuticals (hazard ratio 1.25, 95% confidence interval[CI], 0.60 to 2.61; Table 2) and compared with all non-ICS-users (hazard ratio 1.22, 95% CI, 0.77 to 1.94; Table 2). For influenza patients, the 30-day hazard of admission to ICU was increased among ICS users compared with users of other inhaled pharmaceuticals (hazard ratio 1.43, 95% CI 0.89 to 2.30; Table 2), although not statistically significantly so. No large difference was observed among influenza patients when comparing ICS users with non-ICS users (hazard ratio 1.11, 95% CI 0.85 to 1.46; Table 2).

The 30-day hazard of death among COVID-19 patients was similar among ICS users compared with users of other inhaled pharmaceuticals (hazard ratio 0.84, 95% CI, 0.54 to 1.31; Table 2) and compared with non-ICS users (hazard ratio 1.05, 95% CI, 0.75 to 1.47; Table 2). For influenza patients, the 30-day hazard of death was similar among ICS users compared with users of other inhaled pharmaceuticals (hazard ratio 0.80, 95% CI, 0.63 to 1.01; Table 2) and compared with non-ICS-users (hazard ratio 1.03, 95% CI, 0.87 to 1.22; Table 2). Furthermore, examining the 30-day hazard of mechanical ventilation or death, rather than admission to ICU or death, did not lead to different findings (Table S2).

**Table 2.** Hazard ratio of admission to intensive care unit (ICU) or death within 30 days, among patients with inhaled corticosteroid (ICS) use compared with patients without use of ICS, but use of inhaled β2-receptor agonist and/or muscarinic receptor antagonists, and compared with all patients without ICS use, in hospitalized test-positive individuals for COVID-19 (year 2020) or influenza (year 2010-2018), respectively.

| ICS use comparison group | 30-day relative risk of ICU admission |  | 30-day relative risk of death |  |
| --- | --- | --- | --- | --- |
|  | hazard ratio (95% CI) |  | hazard ratio (95% CI) |  |
|  | Crude* | Adjusted† | Crude* | Adjusted† |
| *During COVID-19 epidemic‡* |  |  |  |  |
| Patients with non-ICS inhaler use | 1.23 (0.60-2.53) | 1.25 (0.60-2.61) | 0.88 (0.57-1.35) | 0.84 (0.54-1.31) |
| All patients without ICS use | 1.15 (0.73-1.79) | 1.22 (0.77-1.94) | 1.20 (0.87-1.65) | 1.05 (0.75-1.47) |
| *During influenza epidemics§* |  |  |  |  |
| Patients with non-ICS inhaler use | 1.52 (0.95-2.43) | 1.43 (0.89-2.30) | 0.84 (0.66-1.06) | 0.80 (0.63-1.01) |
| All patients without ICS use | 1.22 (0.95-1.57) | 1.11 (0.85-1.46) | 1.24 (1.06-1.45) | 1.03 (0.87-1.22) |

\* Adjusted only for age and sex.
† Furthermore adjusted for β2-receptor agonist use and muscarinic receptor antagonist use, in addition to the propensity score containing: atrial fibrillation, dementia, heart failure, hypertension, inflammatory bowel disease, malignancy, renal failure, Charlson Comorbidity index score, number of filled prescription within 90 days, and per oral corticosteroid use.
‡ During the COVID-19 epidemic 282 ICU admissions occurred, of which 29 were ICS users, 16 were users of non-ICS inhalers, and 237 were patients without use of inhalers. Among non-ICU patients 357 deaths occurred, of which 43 were ICS users, 42 were users of non-ICS inhalers, and 272 were patients without use of inhalers.
§ During the 2010-2018 influenza epidemics 951 ICU admissions occurred, of which 224 were ICS users, 77 were users of non-ICS inhalers, and 650 were patients without use of inhalers. Among non-ICU patients 915 deaths occurred, of which 199 were ICS users, 111 were users of non-ICS inhalers, and 605 were patients without use of inhalers.

Among all non-hospitalized individuals who tested positive for SARS-CoV-2, the 30-day hazard of hospitalization was increased in ICS users compared to all individuals with no ICS use (hazard ratio 1.49, 95% CI, 1.16 to 1.91; Table S3), but not compared with users of other inhalants. There was no association between ICS use and hospitalization in non-hospitalized individuals who tested positive for influenza, nor were there any associations between ICS use and death in non-hospitalized individuals who tested positive for influenza or SARS-CoV-2 during 2010–2018 and 2020, respectively (Table S3).

Comparing the Nelson-Aalen cumulative hazard of death within 30-days in the COVID-19 cohort by exposure status, there was no noticeable difference between patients with filled prescriptions for inhaled corticosteroids compared with patients without filled prescriptions for inhaled pharmaceuticals (p-value for log-rank test, 0.28), with approximately 20% dying within 30 days (Figure 1A). A different pattern was observed in the influenza cohort where death was slightly more common among patients not exposed to inhaled corticosteroids (p-value for log-rank test, 0.01), but in contrast only approximately 10% died within 30 days (Figure 1B).

**Figure 1.**
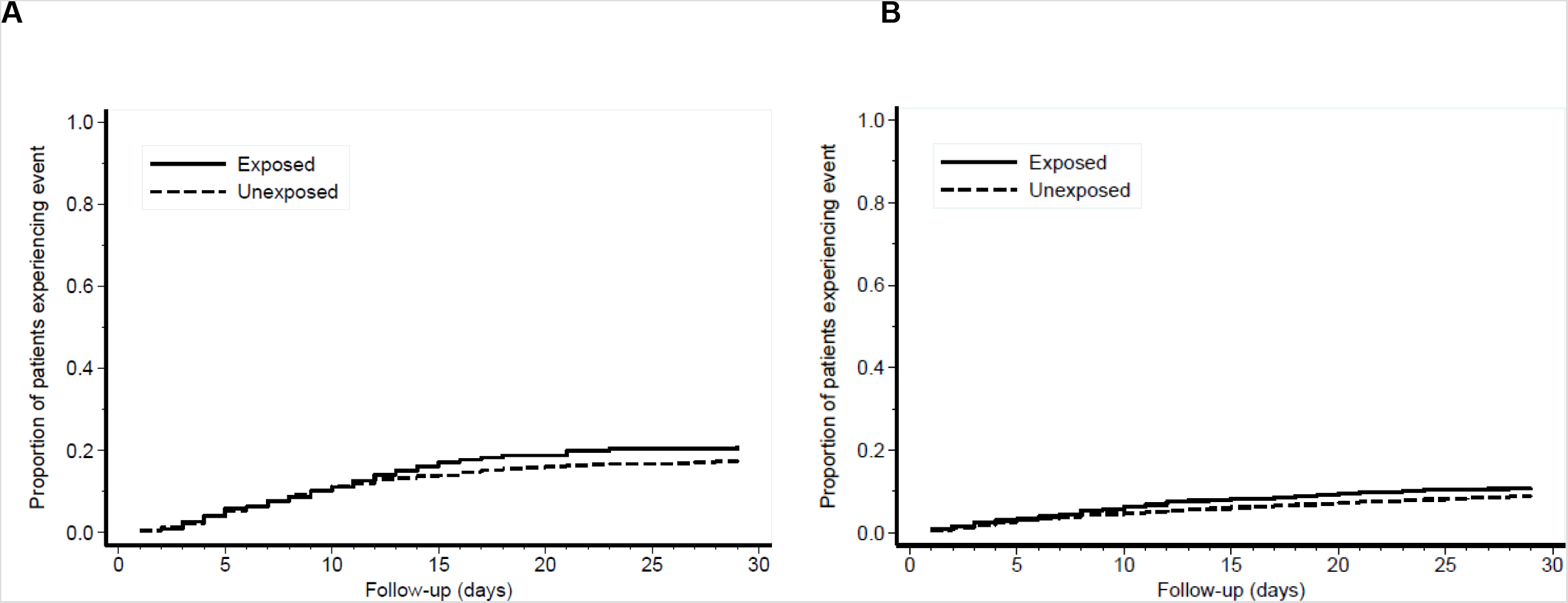
Nelson-Aalen cumulative hazard of death within 30 days, among inhaled corticosteroid (ICS) users compared with non-ICS users, in hospitalized test-positive individuals for SARS-CoV-2 (year 2020, *Panel A*) or influenza (year 2010–2018*, Panel B*), respectively.

Examining the combined 30-day endpoint of admission to ICU and death by subtype of filled ICS, we found no substantial difference between the major subtypes of corticosteroids compared with users of other inhaled pharmaceuticals and non-users of ICS in the COVID-19 cohort (Table S4). However, due to low statistical power we were not able to estimate hazard ratios by prescriptions of beclomethasone, ciclesonide, or mometasone. Nevertheless, comparing the number of COVID-19 patients with prescriptions for ciclesonide and beclomethasone, only seven patients were admitted with prescriptions for ciclesonide, while 17 patients were admitted with prescriptions for beclomethasone, despite yearly beclomethasone use by 0.22% and ciclesonid use by 0.33% of individuals aged 65–79 years in Denmark(16).

In sensitivity analyses, we looked into the effect of inhaled corticosteroid use by excluding individuals with oral corticosteroid use within the last six months (Table S5), different periods of the COVID-19 epidemic (Table S6), different influenza seasons (Table S7), persistency of ICS use (Table S8), and xanthines and leukotriene receptor antagonist use (Table S9), and found no noticeable differences in the hazard of the combined endpoint in different subgroups.

Finally, we investigated the 30-day hazard of death among patients admitted to ICU by use of inhaled corticosteroids (Table S10). In the COVID-19 cohort, we found no statistically significant difference in risk of death among ICU patients by prior usage of inhaled corticosteroids compared with users of other inhaled pharmaceuticals (hazard ratio 0.51 (95% CI, 0.14 to 1.87)) and all non-users of ICS (hazard ratio 0.81 (95% CI, 0.34 to 1.95)), equivalent to the influenza cohort, although the analysis was limited by low statistical power.

## DISCUSSION

In a large nationwide cohort of SARS-CoV-2 test-positive patients, we observed no statistically significant differences in COVID-19 outcomes between patients with inhaled corticosteroid use, patients with other inhaled pharmaceuticals (β_2_-receptor agonist and/or muscarinic receptor antagonists) use, and all patients without inhaled corticosteroid use. Similarly, no consistent effects of inhaled corticosteroids on influenza outcomes during 2010–2018 were observed, which is in line with the results of randomized controlled trials of inhaled corticosteroid use and influenza(17), lending credence to our results on COVID-19. Our findings therefore suggest no major adverse or beneficial effects of inhaled corticosteroid use in COVID-19.

Our results are in agreement with a large, not yet peer-reviewed, cohort study of COPD and asthma patients from the United Kingdom, which reports no increased risk of COVID-19-related death by prescription drug use of inhaled corticosteroids(18). However, the study found a small increased risk of COVID-19-related death by inhaled corticosteroid use, which the authors speculated resulted from unmeasured confounding. Our study did not indicate an increased risk of death with inhaled corticosteroid use, in either crude or adjusted analyses, and benefitted from information on the entire population tested positive for SARS-CoV-2 and subsequently hospitalized (and vice versa), thereby virtually eliminating selection bias. Nevertheless, our supplementary analysis suggested an increased risk of hospital admission among ICS users compared with individuals without use of inhaled pharmaceuticals, but the association disappeared when comparing with all ICS non-users, suggesting confounding by indication.

While our results are reassuring, we cannot rule out minor adverse or beneficial effects of inhaled corticosteroids on COVID-19 outcomes. Moreover, with regard to subtypes of inhaled corticosteroids, we found use of ciclesonide only among seven COVID-19 patients while beclomethasone was used by 17 COVID-19 patients, despite beclomethasone being used approximately 50% more in the Danish population aged 65–79 years. These are small numbers and could be due to differential use by underlying disease severity. However, indications of beneficial effects of ciclesonide, both pre-clinically(6) and in case reports(19,20), argue for further exploration of ciclesonide in randomized controlled trials.

Our cohort study included nationwide information on confirmed cases of SARS-CoV-2 infection and hospitalization in Denmark with negligible loss to follow-up and prospectively registered information on comorbidities and filled prescriptions prior to hospital admission. Furthermore, using hospitalized patients for our main analysis minimized potential selection bias due to potential differential SARS-CoV-2 testing in the general population. Having detailed information on ICS subtypes, persistency of ICS use, and per oral corticosteroid use, additionally allowed us to scrutinize the role of differential inhaled corticosteroids use. Nevertheless, we did not have information on drug dispensation during hospital admission. However, current medications are usually continued upon hospital admission and as nurses dispense medications while in-hospital, we only expect compliance to current medications to be higher during hospitalization.

A further advantage of our study was the ability to compare morbidity of COVID-19 with historical influenza morbidity, as both are viral diseases with principal manifestations in the respiratory system. We found similar effects of inhaled corticosteroid use on COVID-19 morbidity in 2020 and influenza morbidity during 2010–2018. The finding of no effect of inhaled corticosteroids on influenza was consistent with a meta-analysis on randomized controlled trials of inhaled corticosteroid treatment(17). Furthermore, in parallel with other reports we found COVID-19 to be associated with a markedly higher level of overall morbidity and mortality compared with influenza(21,22). This finding underscores the severity of COVID-19, since our COVID-19 cohort was both younger and generally healthier than our influenza cohort was.

Together with the findings of others, our study supports continued use of inhaled corticosteroid according to current guidelines. Although suggesting no major adverse or beneficial effect of inhaled corticosteroids on COVID-19, our study is no substitution for randomized controlled trials of inhaled corticosteroids in the treatment of COVID-19. As also suggested by others(23), inhaled corticosteroids could potentially limit both short-term and long-term COVID-19 morbidity and trials are underway(24). Specifically, one potential avenue to be explored given the results of ours and other studies, is replacing existing inhaled corticosteroid treatment with either ciclesonide or non-ciclesonide corticosteroids at random in patients at risk of COVID-19. However, no inhaled corticosteroids should be used as treatment for COVID-19, beyond existing pharmaceutical use, unless administered as part of a clinical trial.

Taken together, our study found no effect of inhaled corticosteroids on COVID-19 morbidity or mortality compared with either use of non-corticosteroid inhalers or no inhaled corticosteroid use overall. Treatment with inhaled corticosteroids should therefore follow current guidelines, unless investigated in randomized controlled trials under the supervision of medical professionals.

## Data Availability

The data used in the study can be obtained by first submitting a research protocol to the Danish Data Protection Agency(Datatilsynet) and once permission has been received, by applying the Ministry of Healths Research Service(Forskerservice) and Statistics Denmark(Danmarks Statistik) for access to the data. The data do therefore not belong to the authors and they are not permitted to share them, except in aggregate form.

## REFERENCES

1. Zhu N, Zhang D, Wang W, Li X, Yang B, Song J, et al. A Novel Coronavirus from Patients with Pneumonia in China, 2019. N Engl J Med. 2020 Jan 24;

2. Beigel JH, Tomashek KM, Dodd LE, Mehta AK, Zingman BS, Kalil AC, et al. Remdesivir for the Treatment of Covid-19 — Preliminary Report. N Engl J Med. 2020 May 22;

3. Horby P, Lim WS, Emberson JR, Mafham M, Bell JL, Linsell L, et al. Dexamethasone in Hospitalized Patients with Covid-19 — Preliminary Report. N Engl J Med. 2020 Jul 17;NEJMoa2021436.

4. Bhatraju PK, Ghassemieh BJ, Nichols M, Kim R, Jerome KR, Nalla AK, et al. Covid-19 in Critically Ill Patients in the Seattle Region — Case Series. N Engl J Med. 2020 Mar 30;NEJMoa2004500.

5. Peters MC, Sajuthi S, Deford P, Christenson S, Rios CL, Montgomery MT, et al. COVID-19 Related Genes in Sputum Cells in Asthma: Relationship to Demographic Features and Corticosteroids. Am J Respir Crit Care Med. 2020 Apr 29;

6. Jeon S, Ko M, Lee J, Choi I, Byun SY, Park S, et al. Identification of antiviral drug candidates against SARS-CoV-2 from FDA-approved drugs. Antimicrob Agents Chemother. 2020 Jun 23;64(7).

7. Lippi G, Henry BM. Chronic obstructive pulmonary disease is associated with severe coronavirus disease 2019 (COVID-19). Respir Med. 2020 Mar;105941.

8. Williamson EJ, Walker AJ, Bhaskaran K, Bacon S, Bates C, Morton CE, et al. OpenSAFELY: factors associated with COVID-19 death in 17 million patients. Nature. 2020 Jul 8;1–11.

9. Pedersen CB, Gøtzsche H, Møller JO, Mortensen PB. The Danish Civil Registration System. A cohort of eight million persons. Dan Med Bull. 2006 Nov;: 53(4): 441–9.

10. Voldstedlund M, Haarh M, Mølbak K, the MiBa Board of Representatives C. The Danish Microbiology Database (MiBa) 2010 to 2013. Eurosurveillance. 2014 Jan 9;: 19(1): 20667.

11. Voldstedlund M, Haahr M, Emborg H-D, Bang H, Krause T. Real-time surveillance of laboratory confirmed influenza based on the Danish microbiology database (MiBa). Stud Health Technol Inform. 2013; 192: 978.

12. Pottegård A, Kristensen KB, Reilev M, Lund LC, Ernst MT, Hallas J, et al. Existing data sources in clinical epidemiology: The danish covid-19 cohort. Clin Epidemiol. 2020 Aug 12; 12: 875–81.

13. Schmidt M, Schmidt SAJ, Sandegaard JL, Ehrenstein V, Pedersen L, Sørensen HT. The Danish National Patient Registry: a review of content, data quality, and research potential. Clin Epidemiol. 2015; 7: 449–90.

14. Pottegård A, Schmidt SAJ, Wallach-Kildemoes H, Sørensen HT, Hallas J, Schmidt M. Data Resource Profile: The Danish National Prescription Registry. Int J Epidemiol. 2016 Oct 27; 25(3): dyw213.

15. Helweg-Larsen K. The Danish register of causes of death. Scand J Public Health. 2011 Jul 20; 39(7): 26–9.

16. Sundhedsdatastyrelsen – Statistikker [Internet]. [cited 2020 Aug 4]. Available from: https://medstat.dk/en

17. Dong YH, Chang CH, Wu FLL, Shen LJ, Calverley PMA, Löfdahl CG, et al. Use of inhaled corticosteroids in patients with COPD and the risk of TB and influenza: A systematic review and meta-analysis of randomized controlled trials. Chest. 2014 Jun 1; 145(6): 1286–97.

18. Schultze A, Walker AJ, MacKenna B, Morton CE, Bhaskaran K, Brown JP, et al. Inhaled corticosteroid use and risk COVID-19 related death among 966,461 patients with COPD or asthma: an OpenSAFELY analysis. medRxiv. 2020 Jun 20;2020.06.19.20135491.

19. Nakajima K, Ogawa F, Sakai K, Uchiyama M, Oyama Y, Kato H, et al. A Case of Coronavirus Disease 2019 Treated With Ciclesonide. Vol. 95, Mayo Clinic Proceedings. Elsevier Ltd; 2020. p. 1296–7.

20. Iwabuchi K, Yoshie K, Kurakami Y, Takahashi K, Kato Y, Morishima T. Therapeutic potential of ciclesonide inahalation for COVID-19 pneumonia: Report of three cases. J Infect Chemother. 2020 Jun 1;: 26(6): 625–32.

21. Faust JS, Del Rio C. Assessment of Deaths from COVID-19 and from Seasonal Influenza. Vol. 180, JAMA Internal Medicine. American Medical Association; 2020. p. 1045–6.

22. Nersesjan V, Amiri M, Christensen H, Benros ME, Kondziella D. 30-day mortality and morbidity in COVID-19 versus influenza: A population-based study. medRxiv. 2020 Jul 28;2020.07.25.20162156.

23. Nicolau D V, Bafadhel M. Inhaled corticosteroids in virus pandemics: a treatment for COVID-19? Lancet Respir Med. 2020 Jul;0(0).

24. Home – ClinicalTrials.gov [Internet]. [cited 2020 Aug 7]. Available from: https://clinicaltrials.gov/

